# Analysis of electronic health records reveals medication-related interference on point-of-care urine drug screening assays

**DOI:** 10.1101/2020.04.03.20052647

**Authors:** Nadia Ayala-Lopez, Jennifer M. Colby, Jacob J. Hughey

## Abstract

**Background:** Point-of-care (POC) urine drug screening (UDS) assays provide immediate information for patient management. However, POC UDS assays can produce false positive results, which may not be recognized until confirmatory testing is completed several days later. To minimize the potential for patient harm, it is critical to identify sources of interference. Here we applied an approach based on statistical analysis of electronic health record (EHR) data to identify medications that may cause false positives on POC UDS assays.

**Methods:** From our institution’s EHR data, we extracted 120,670 POC UDS and confirmation results, covering 12 classes of target drugs, along with each individual’s prior medication exposures. For a given assay and medication ingredient, we quantified potential interference as an odds ratio from logistic regression. We evaluated interference experimentally by spiking compounds into drug-free urine and testing the spiked samples on the POC device (Integrated E-Z Split Key Cup II, Alere).

**Results:** Our dataset included 446 false positive UDS results (presumptive positive screen followed by negative confirmation). We quantified potential interference for 528 assay-ingredient pairs. Of the six assay-ingredient pairs we evaluated experimentally, two showed interference capable of producing a presumptive positive: labetalol on the MDMA assay (at 200 μg/mL) and ranitidine on the methamphetamine assay (at 50 μg/mL). Ranitidine also produced a presumptive positive for opiates at 1600 μg/mL and for propoxyphene at 800 μg/mL.

**Conclusions:** These findings support the generalizability of our approach to use EHR data to identify medications that interfere with clinical immunoassays.

## Introduction

Urine drug screens (UDS) are commonly based on immunoassays, which are sensitive and cost-effective. As with nearly any laboratory test, however, immunoassays are susceptible to interference by medications, vitamins, and other substances. Because many UDS assays are designed to recognize multiple related substances, they may be particularly susceptible to interference from structurally similar compounds [1]. Due to these assays’ relatively low specificity, positive UDS results are considered presumptive. Distinguishing between true positive and false positive UDS results requires confirmatory testing based on mass spectrometry.

Point-of-care (POC) UDS immunoassays are designed to be used at or near the site of patient care, which enables immediate incorporation of the results into patient management. Like the immunoassays performed in a clinical laboratory, POC UDS assays are susceptible to interference from medications. Given that POC results are acted on before confirmation results are available, providers are often cautioned about possible interference and false positives. To educate providers and minimize the potential for patient harm, many laboratories compile lists of interfering substances based on information from the manufacturer, the scientific literature, and experience. Unfortunately, these lists tend to be limited in scope, raising the possibility that multiple sources of interference remain unknown.

We recently developed and validated an approach to identify medications that may interfere with laboratory-based UDS assays [2]. Here we adapted our approach to search for medications that interfere with POC UDS assays. Our statistical analysis of electronic health record (EHR) data used more than 120,000 POC UDS results generated over a five-year period. We then experimentally validated predicted interferences through spiking studies.

## Materials and Methods

Code and summary results for this study are available at https://doi.org/10.6084/m9.figshare.12067401. The Vanderbilt Institutional Review Board reviewed and approved this study as non-human subjects research (IRB# 081418 and 190165).

### Extraction of POC UDS results and drug exposures from electronic health record data

EHR data came from the Synthetic Derivative, a collection of deidentified clinical data from Vanderbilt University Medical Center (VUMC) [3]. We followed the same strategy as our previous study [2] to collect UDS results related to the point-of-care test cup (Integrated E-Z Split Key Cup II, Alere) and the corresponding reflexed confirmation results. The confirmation assays are laboratory-developed tests based on GC-MS (amphetamine, barbiturates, cannabinoids, cocaine metabolite, MDMA, methadone, methamphetamine, opiates, oxycodone, propoxyphene) or LC-MS/MS (benzodiazepines and buprenorphine). If a presumptive positive UDS result was accompanied by a positive or negative confirmation, we denoted it as a true positive or false positive, respectively.

As in our previous study, for each person in the dataset, we identified drug exposures documented between 1 and 30 days prior to each UDS result. We excluded UDS results that occurred less than 30 days after the person’s first ever visit at VUMC, since we would lack a prior 30 days of documented drug exposures. Documented drug exposures are available as structured data in the Synthetic Derivative and come primarily from medication lists. We mapped each drug to its active ingredient(s) using RxNorm [4]. As described previously, having a documented exposure within 30 days is only a proxy for being exposed at the time of the UDS [2].

### Statistical analysis of drug exposures and UDS results

We quantified associations between drug exposures and false positive UDS results similarly to in our previous study. For an assay-ingredient pair, we fit a logistic regression model in which the dependent variable corresponded to the UDS result (negative or false positive) and the independent variable corresponded to presence or absence of prior exposure to the ingredient. We fit each model using Firth’s method [19,20], which yielded a log odds ratio and standard error. We then used an Empirical Bayes approach called adaptive shrinkage [5] to estimate the posterior mean of the log odds ratio and the corresponding 95% credible interval for each assay-ingredient pair. These are the quantities we use throughout the paper. We fit a model for an assay-ingredient pair if at least two individuals exposed to the ingredient had a false positive result on the assay. We quantified associations between ingredients and true positive UDS results in the same way. To distinguish the effects of concurrent exposure to multiple ingredients, we fit a logistic regression model with a term for each ingredient of interest. We defined known interferents as substances whose ability to cause a presumptive positive was described in the assay’s package insert.

### Experimental validation of interference

For each selected compound, we spiked a reference standard into drug-free urine at various concentrations and tested the spiked urine samples in the Integrated E-Z Split Key Cup II (Alere San Diego, Inc., San Diego, CA). We purchased reference standards from Sigma-Aldrich (Milwaukee, WI), Tocris Biosciences (Bristol, UK), and Santa Cruz Biotechnology (Dallas, TX). We prepared stock solutions of each standard in 80% DMSO in water (labetalol, meloxicam, and ranitidine), 100% DMSO (5’-carboxy meloxicam), 80% methanol in water (prazosin), and 100% methanol in water (furosemide). We spiked the urine samples using a fixed volume of 20% spiking solution, made of a combination of diluent and stock solution, including one sample per compound with only diluent to serve as a negative control. In most cases, we tested the maximum technically feasible concentration for a compound, given the limits of solubility, the concentration of the reference material, and the fixed 20% spiking volume. We performed positive controls using Liquicheck Urine Toxicology Control Level C4 (Bio-Rad, Hercules, CA) and high calibrators from Immunalysis (Pomona, CA). The POC assays provide qualitative results that are interpreted visually, making the experimental validation somewhat subjective.

## Results

We first assembled a dataset of UDS and confirmation results related to our institution’s point-of-care device, which includes immunoassays for 12 classes of target drugs (Table 1). The dataset included 120,670 results from 1,163 individuals, along with each person’s documented drug exposures occurring between 1 and 30 days prior. Each UDS result was preceded by exposure to a median of 3 ingredients and mean of 6.8 ingredients. The assay with by far the highest true positive rate was for buprenorphine, consistent with the POC device being used primarily as part of our institution’s medication assisted treatment program.

**Table 1.**
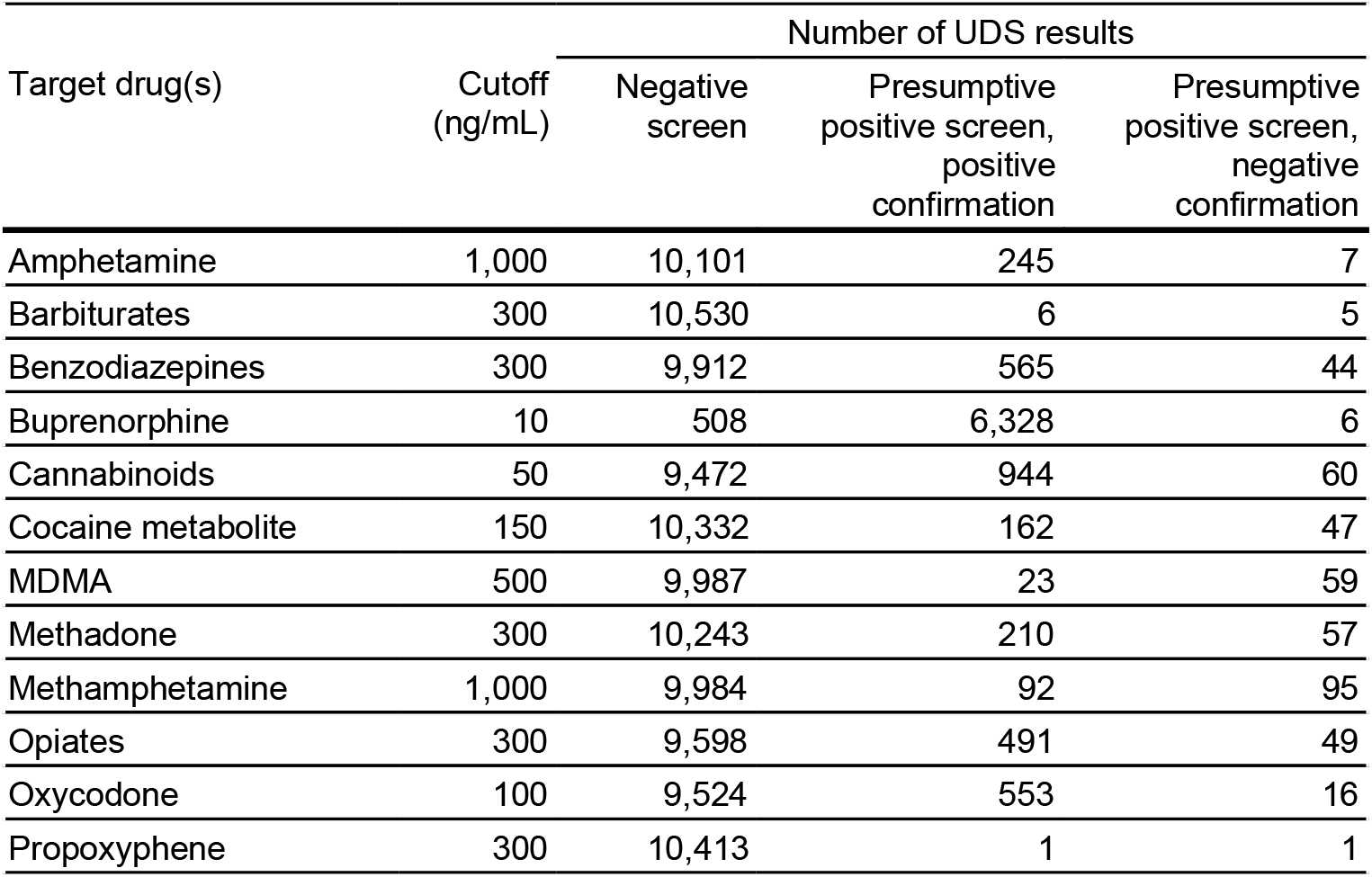
Characteristics of urine drug screening immunoassays in this study.

Similarly to our previous study [2], we used the dataset to calculate two types of odds ratios: one for the association with false positive UDS results (which we call OR_FP_) and one for the association with true positive UDS results (which we call OR_TP_). As before, we hypothesized that OR_FP_ would identify potential interferents on a given assay and OR_TP_ would identify assay targets (thereby serving as a positive control). Altogether, we calculated OR_FP_ for 528 assay-ingredient pairs and OR_TP_ for 1,796 assay-ingredient pairs (Supplemental Tables 1 and 2).

Because of the relatively low counts of false positive UDS results, we could only calculate OR_FP_ for one known interferent. However, most assay targets had among the highest OR_TP_ on their respective assay (Figure 1), indicating that even on this smaller dataset, our approach detects the effects of drug exposure on UDS results.

**Figure 1.**
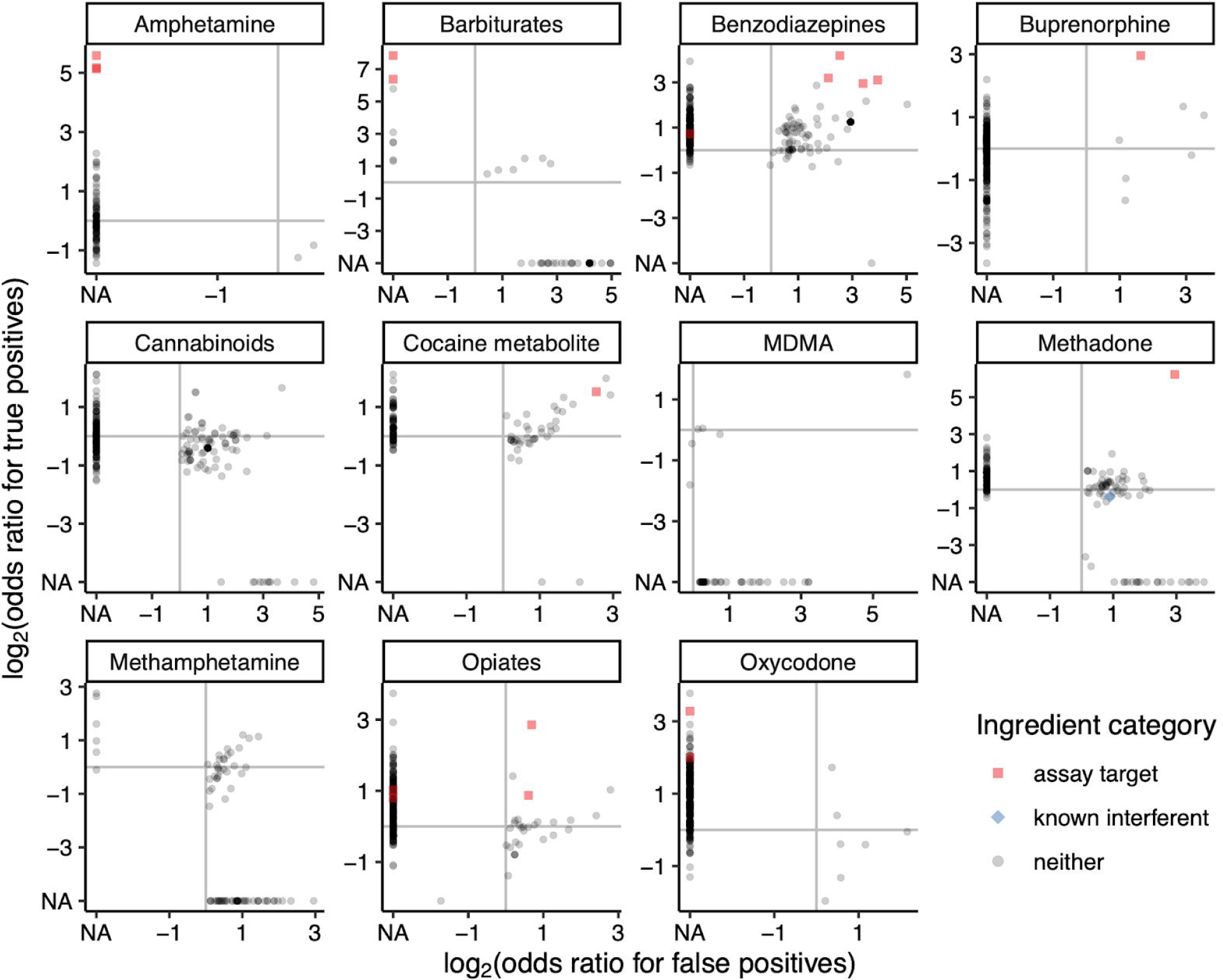
Associations with false positive and true positive UDS results for all tested assay-ingredient pairs. Each plot corresponds to an assay, and each point corresponds to an ingredient. A log2 odds ratio of NA indicates that the association was not tested, as fewer than two individuals had a false positive (NA on x-axis) or true positive (NA on y-axis) UDS result preceded by exposure to the given ingredient. Only one known interferent was tested for any association, and it is on the methadone assay.

We selected the most promising potentially interfering ingredients for experimental validation (Table 2), although we expected that because of the low counts fewer of them would actually produce a presumptive positive. We selected meloxicam and furosemide for validation on the cannabinoids assay based on multivariate regression (Supplemental Table 3).

**Table 2.** Strongest associations with false positive UDS results, which were selected for experimental evaluation. Numbers of UDS results correspond to those preceded by exposure to the given ingredient. NA indicates the compound did not cause a presumptive positive up to the highest concentration tested. ^a^Neither meloxicam nor 5’-carboxy meloxicam caused a presumptive positive.

| Assay | Ingredient | Number of UDS results |  |  | Odds ratio (OR<sub>FP</sub>) | 95% credible interval |  | Conc. causing a presumptive positive (µg/mL) |
| --- | --- | --- | --- | --- | --- | --- | --- | --- |
|  |  | Negative | True positive | False positive |  | Lower | Upper |  |
| Cannabinoids | Meloxicam | 5 | 0 | 2 | 28.1 | 7.1 | 78.0 | NA<sup>a</sup> |
| Cannabinoids | Furosemide | 101 | 1 | 7 | 11.4 | 5.4 | 22.0 | NA |
| MDMA | Labetalol | 143 | 2 | 30 | 61.9 | 39.9 | 80.5 | 200 |
| Methadone | Labetalol | 171 | 5 | 9 | 10.4 | 5.3 | 20.5 | NA |
| Methadone | Prazosin | 198 | 1 | 9 | 9.0 | 4.6 | 18.5 | NA |
| Methamphetamine | Ranitidine | 97 | 0 | 7 | 7.8 | 3.6 | 17.0 | 50 |

Of the six assay-ingredient pairs we evaluated (Supplemental Figure 1), two showed interference capable of producing a presumptive positive: labetalol on the MDMA assay and ranitidine on the methamphetamine assay (Table 2). As incidental findings, ranitidine also produced a presumptive positive for opiates at 1600 μg/mL and for propoxyphene at 800 μg/mL.

## Discussion

Knowledge of substances that can cause spurious results is critical for point-of-care testing. Here we applied a statistical approach to detect sources of interference on POC urine drug screening assays.

The current study differs from our previous work in several important ways. First, the current dataset included ∼6-fold fewer UDS results overall and ∼16-fold fewer false positives. The latter is at least partly because, compared to the laboratory-based assays, the POC assays have higher cutoff concentrations for presumptive positive results. Second, because the POC assays at our institution are used primarily in outpatient medication assisted treatment clinics, the current dataset also included ∼35-fold fewer unique patients. All these factors likely contributed to why we identified fewer interfering medications in the current study than in the previous study.

An important future step, especially relevant to testing at the point of care, is to integrate findings from this study and others into the EHR so that providers are automatically notified when a result is likely spurious. Overall, the repeated success of our approach supports its generalizability, and hints at the broader potential for analysis of EHR data to advance laboratory medicine.

## Data Availability

Code and summary results for this study are available at https://doi.org/10.6084/m9.figshare.12067401.

## Acknowledgments

None declared.

## Author contributions

All the authors have accepted responsibility for the entire content of this submitted manuscript and approved submission.

## Research funding

This work was supported in part by CTSA award UL1TR002243 from NCATS/NIH and the Vanderbilt Institute for Clinical and Translational Research (VICTR) grant VR54098. The Vanderbilt Synthetic Derivative is supported by institutional funding and by CTSA award UL1TR002243 from NCATS/NIH.

## Employment or leadership

None declared.

## Honorarium

None declared.

